# Applying Machine-Learning and Deep-Learning to Predict Depression from Brain MRI and Identify Depression-Related Brain Biology

**DOI:** 10.1101/2025.02.14.25322326

**Authors:** Jiayue-Clara Jiang, Camille Brianceau, Elise Delzant, Romain Colle, Hugo Bottemanne, Emmanuelle Corruble, Naomi R Wray, Olivier Colliot, Sonia Shah, Baptiste Couvy-Duchesne

**Affiliations:** Institute for Molecular Bioscience, The University of Queensland, St Lucia, QLD, Australia; Sorbonne University, Paris Brain Institute - ICM, CNRS, Inria, Inserm, AP-HP, Hôpital de la Pitié Salpêtrière, F-75013, Paris, France; Service Hospitalo-Universitaire de Psychiatrie de Bicêtre, Mood Center Paris Saclay, Assistance Publique-Hôpitaux de Paris, Hôpitaux Universitaires Paris-Saclay, Hôpital de Bicêtre, Le Kremlin Bicêtre, F-94275, France; MOODS Team, INSERM 1018, CESP (Centre de Recherche en Epidémiologie et Santé des Populations), Université Paris-Saclay, Faculté de Médecine Paris-Saclay, Le Kremlin Bicêtre, F-94275, France; Department of Psychiatry, University of Oxford, Oxford

## Abstract

The accuracy of grey-matter predictors of depression has remained limited. In this study, brain-based predictors of major depressive disorder (MDD) were trained using machine-learning (Best Linear Unbiased Predictors [BLUP]) and deep-learning (ResNet3D) techniques applied to high-dimensional (voxel-wise) grey-matter structure extracted from T1-weighted structural MRI. The training sample comprised 987 MDD cases and 3,934 controls from the UK Biobank. Predictors were evaluated in an independent sub-cohort of 483 MDD cases and 1,939 controls from the UK Biobank and replicated in a clinical cohort (DEP-ARREST CLIN) of 64 cases and 32 controls. In the UK Biobank, logistic regression showed a significant association between the BLUP predictor and MDD status (AUC=0.57; OR=1.28 [1.15-1.43]; p-value=1.1×10^−5^), which was confirmed in both males and females. By partitioning the BLUP predictor by brain regions of interest (ROI), we found nominal significance supporting the contribution of previously identified MDD-related ROIs (e.g. hippocampus and amygdala). The BLUP predictor overlapped partially with a polygenic score (PGS) of major depression but also captured a signal that was not captured by the genetic score (combined AUC=0.66, p-value=0.024 when compared to PGS alone). No association passed multiple testing correction in the DEP-ARREST CLIN cohort, likely due to the small sample size. In contrast, the deep-learning predictor did not show a significant association with MDD after multiple testing corrections. Our novel application of the BLUP method shows promising predicting accuracy and suggests new leads to overcome the remaining challenges in predicting MDD from brain imaging.

## Introduction

Despite being a leading contributor to the global burden of disease for people aged 10 to 74 years [1], depression remains poorly understood, possibly due to its heterogeneous presentation and multi-causal aetiology influenced by genetic, environmental and social factors [2]. Depression genetic risk loci identified from genome-wide association studies (GWAS) have been shown to be enriched for genes expressed in specific brain regions, specifically the anterior cingulate cortex and frontal cortex [3]. Furthermore, several brain structural features, such as larger hippocampal volumes and higher frontal cortical thickness, have been found to predict remission after antidepressant treatment [4]. These pieces of evidence converge to suggest a complex and dynamic implication of brain regions in leading to depression and over the course of depression episodes and treatment.

Brain imaging, such as Magnetic Resonance Imaging (MRI), has advanced the understanding of the brain structures and functions associated with depression. Historically, neuroimaging studies have been limited by small sample sizes, leading to few robust results [5, 6]. The Enhancing NeuroImaging Genetics through Meta-Analysis (ENIGMA) consortium has gathered brain images from thousands of depression cases and controls from more than 20 sites to perform large-scale analyses [7]. They consistently reported a reduced hippocampal volume amongst individuals with depression, whereas the findings on structural changes in other brain regions, such as the amygdala and thalamus, have remained conflicting [2]. The variability in results may be due to different imaging protocols and machines as well as the heterogeneous nature of depression, manifesting as different symptoms, severity and subtype categories. At the same time, the ENIGMA-led efforts to build brain-based predictors of depression have failed, highlighting the difficulty of this task [8]. More recently, the Depression Imaging REsearch ConsorTium (DIRECT) has collected structural and resting-state functional MRI (rsfMRI) data from 2,428 participants, and provided insights into changes in the functional connectivity in patients with depression, though they have not attempted to build predictors of depression status [9].

Another limitation of previous brain imaging studies of depression is that they considered high-level brain measurements, such as cortical thickness and subcortical volumes in regions of interests (ROIs) defined using brain atlases [10]. While this reduces multiple testing and computation, these high-level measurements lack granularity and may fail at detecting subtle and localised associations in the brain [11, 12]. This may explain why approaches that relied on data reduction techniques have failed to yield depression predictors with detectable prediction accuracy [13].

In this project, we aimed to train predictors of major depressive disorders (MDD) on pre-processed high-dimensional (voxel-based) brain MRI data from the UK Biobank [10], using machine-learning and deep-learning algorithms, which can handle high-dimensional data [5, 14]. Although the UK Biobank was not focused on depression, more than a thousand participants with brain MRI data available had a recorded episode of MDD. The large sample and homogeneous image acquisition (compared to the data gathered by consortia) may facilitate the training of algorithms. Our pre-processing includes the normalisation of images into a standard space, which should simplify the learning task and ensure that both deep-learning and machine-learning can be trained on the same images. Lastly, the voxel-based representation of grey matter retains more granularity and signal of interest than a ROI-based approach. To progress in understanding depression-related brain biology, we sought to identify the brain regions contributing to the algorithms’ predictive ability. Lastly, we evaluated if the brain-based predicted risk captured independent information from a genetic predictor of depression.

## Materials and Methods

An overview of study is presented in Supplemental Figure 1.

### UK Biobank

The UK Biobank cohort consists of over 500,000 volunteers (aged 40-69 years at baseline) from the UK, recruited between 2006 and 2010 [15]. We used the UK Biobank data collected between 2006 and March 2023, and excluded individuals who withdrew from the study. At the time of analysis, the brain images for 39,994 participants were available [16]. After excluding individuals who did not answer or selected “prefer not to answer” for the smoking status question, or lacked information on BMI at baseline, a total of 39,740 individuals were retained for further analysis.

### MDD status and antidepressant use ascertainment in the UK Biobank

The full list of UK Biobank data-fields and entry codes used to define MDD cases are summarised in Supplemental Table 1. Briefly, MDD cases were defined by ICD-10 (data-field 41270) or ICD-9 (data-field 41271) codes for depressive episode, recurrent depressive disorder, persistent mood (affective) disorders, other mood (affective) disorders or unknown mood (affective) disorders. We excluded MDD cases with ICD-10 or ICD-9 codes of bipolar disorder or schizophrenia, had self-reported schizophrenia (data-field 20002), had an ICD-10 code of multiple personality disorder, or were taking antipsychotics (data-field 20003). The screened controls were defined as individuals who had no diagnosis of MDD, bipolar disorder or schizophrenia, had no ICD diagnosis of personality or neurotic disorders, had no self-reported history of mood disorders, and were not on any antidepressants or antipsychotic medications. Amongst individuals who had MRI data available and with complete records of age at baseline (calculated using data-field 34 “year of birth” and data-field 53 “Date of attending assessment centre”), sex (data-field 31), BMI (data-field 21001) and smoking status (data-field 20116) at baseline, we identified a total of 1,496 MDD cases and 27,741 screened controls. To evaluate the potential effects of antidepressants on brain structure, we also identified individuals who self-reported to have used antidepressants at the initial or follow-up assessments.

### Training and test cohorts

To construct the training cohort, we randomly selected 1,000 MDD cases. Using the “MatchIt” R package (version 4.5.5), we selected four controls for each MDD case, matched by sex, genetic ancestry (ancestry calling is described in Yengo et al. [17]), age at baseline, smoking status (ever smoked or never smoked) at baseline, and BMI at baseline. The test cohort consisted of the remaining MDD cases (N=496) and matched controls not included in the training sample. After excluding individuals whose brain MRI images were deemed “non-usable” as previously described [16], the training cohort consisted of 987 MDD cases and 3,934 controls, and the test cohort consisted of 483 MDD cases and 1,939 controls (demographic summary is presented in Supplemental Table 2).

### Pre-processing of brain MRI images

Using the T1-volume pipeline from the Clinica software platform [18], we processed the T1-weighted MRI images from the UK Biobank with the statistical parametric mapping (SPM) software package [19]. Tissue segmentation, bias correction and spatial normalisation were performed, and the grey matter in the brain images was extracted. A Dartel template was generated from the images of 1,000 randomly selected study subjects from the training cohort, and the brain MRI images of study subjects were normalised and aligned to a common space by projection onto the Dartel template. This step ensured voxel-wise correspondence across the subjects. 2,122,945 voxels per image were included in subsequent analysis, though only a fraction corresponds to the grey matter.

### Brain atlas for interpretation

Voxels were mapped to brain anatomical regions using the cerebellum atlas by Diedrichsen et al. [20], and the cortical and subcortical atlases by Makris et al. [21], Frazier et al. [22] and Goldstein et al. [23].

### Training a machine-learning predictor

We used the OmicS-data-based Complex trait Analysis (OSCA) tool [24] to train a machine-learning-based predictor of MDD risk using the Best Linear Unbiased Prediction (BLUP) [25]. BLUP is a statistical framework and a special case of machine-learning algorithms that maximises the restricted maximum likelihood (REML). As such, it does not require cross-validation and is computationally efficient [26].

Voxels with 0 values for most-to-all individuals are on the edge or outside of the grey matter and cannot be fitted in a linear model. Hence, we excluded voxels with mean intensity value < 0.1 and variance < 0.01, leaving a total of 415,836 voxels from 150 ROIs for the BLUP analysis. The joint, conditional effects of the voxels were estimated from the training cohort using REML, and the voxel weights were applied to the test cohort to generate the BLUP predictor.

In the training cohort, we also estimated the proportion of phenotypic variance captured by all voxel-wise intensities (previously coined “morphometricity” [27]) using OSCA [24]. We tested the statistical significance of a non-zero morphometricity using a likelihood ratio test [24]. The morphometricity model included as covariates age at baseline, age at imaging, sex, genetic ancestry, smoking status and BMI at baseline, as well as a number of MRI imaging covariates, namely mean rfMRI head motion (data-field 25741), scanner lateral (X) brain position (data-field 25756), scanner transverse (Y) brain position (data-field 25757), scanner longitudinal (Z) brain position (data-field 25758), intensity scaling for T1 (data-field 25925), volume of EstimatedTotalIntraCranial (whole brain) (data-field 26521), inverted signal-to-noise ratio in T1 (data-field 25734), and inverted contrast-to-noise ratio in T1 (data-field 25735).

### Training a deep-learning predictor

We trained a deep-learning model to perform a classification task that predicts MDD status using the ClinicaDL software platform [28]. Briefly, tensors were extracted from the MRI images to prepare for downstream training that relies on the PyTorch deep-learning library [29]. The training cohort was split into a training and validation set at a 3:1 ratio, while balancing age at baseline, age at imaging, sex, genetic ancestry, smoking status and BMI at baseline, and birth year. We trained a ResNet3D architecture using a weighted sampler (giving a stronger weight to underrepresented class), 50 epochs and a 10% dropout rate. The model generated in the epoch with the lowest cross-entropy loss on the validation set was selected as the final model for prediction in the test cohort (hereby referred to as the deep-learning predictor).

### Evaluating prediction accuracy

We compared the predicted scores between the true MDD cases and controls in the test cohort using t-tests to evaluate the prediction accuracy of the BLUP and deep-learning predictors. We also investigated the covariate-adjusted association between the predictors and MDD status using logistic regression, where the predictors were normalised to have a mean of 0 and a standard deviation (SD) of 1. The logistic regression model adjusted for age at baseline, age at imaging, sex (except in sex-stratified analyses), genetic ancestry, and smoking status and BMI at baseline. We reported the odds ratios (OR) from logistic regression. Given the overrepresentation of female cases and controls in our training cohort, we repeated the logistic regression analysis in males and females separately. We used a two-sided p-value < 0.0125 (multiple testing correction for two predictors and two sexes) to declare statistical significance.

We performed sensitivity analysis where the logistic regression model included MRI imaging covariates as additional covariates. As only 61.3% of the MDD cases in the test cohort had self-reported antidepressant use, we repeated the logistic regression adjusting for antidepressant use. We performed additional sensitivity analyses after excluding individuals with a recorded MDD episode (estimated using the date of the ICD-9 and ICD-10 records) more than 5 years prior to or after imaging.

Using the “roc” function from the pROC [30] R package (version 1.18.0), we also quantified prediction accuracy using the area under the receiver operator characteristic curve (AUC), which can be interpreted as a probability of a case having a higher predicted score than a control. The AUC of different predictors (e.g. BLUP versus deep-learning) were compared using DeLong’s test for two correlated ROC curves, performed with the “roc.test” function from pROC [30].

### Identifying MDD-associated ROIs from the BLUP predictor

We built ROI-specific predictors by restricting the score to the voxels in each ROI (using BLUP weights estimated across the whole brain, as in the previous analysis). We used logistic regression to investigate the association between each ROI-specific BLUP predictor and MDD risk, while adjusting for covariates as in the main analysis. A p-value < 0.00033 (Bonferroni correction for 150 ROIs) was used to declare statistical significance.

### Investigating the association with genetically predicted MD risks

GWASs have identified many genetic variants implicated in the risk of depression [3, 31, 32]. As an estimate for the lifetime genetic liability of an individual to depression, we developed polygenic scores (PGS) for major depression (MD, different from MDD as MD includes self-reported cases). We used SBayesR [33] with the banded LD matrix reference to derive the per-allele weights from the latest MD GWAS [32], after the exclusion of non-European-ancestry participants and participants from UK Biobank and 23andme.

As the SNP weights were estimated from GWAS summary statistics derived from European-ancestry individuals, we calculated the PGS_MD_ for the subset of individuals of European ancestry in the UK Biobank test cohort using PLINK1.9 [34].

Using linear regression, we estimated the correlation of PGS_MD_ with the BLUP and deep-learning predictors (scaled to a mean of 0 and a SD of 1), adjusting for sex (except in sex-stratified analysis), age at baseline, age at imaging, smoking and BMI at baseline and 10 genetic principal components (PC). A two-sided p-value < 0.0125 (Bonferroni correction for two predictors and two sexes) was used to declare statistical significance.

### Replication in the DEP-ARREST CLIN cohort

We sought to replicate our results in the DEP-ARREST CLIN cohort. The DEP-ARREST CLIN cohort consisted of 64 patients and 32 controls aged 18-65 years old. The cases were hospitalised in the Department of Psychiatry of Bicêtre University Hospital with a diagnosis of current Major Depressive Episode (MDE) in the context of MDD (DSM-IVTR) based on the Mini International Neuropsychiatric Interview (MINI) [35]. Patients with a Hamilton Depression Rating Scale 17 items (HDRS) score ≥ 18 were included, and were excluded if they had brain syndromes, unstable medical conditions, bipolar disorders (DSM-IVTR) or current treatment with mood stabilisers, psychotic disorders (DSM-IVTR) or current treatment with antipsychotics, current substance abuse or dependence (DSM-IVTR), as well as pregnancy, breastfeeding, and contra-indications to cerebral MRI. All patients were systematically assessed by a psychiatrist.

We processed the brain MRI images using Clinica as per the UK Biobank. Images were normalised to the DARTEL template created from the UK Biobank images. We applied the BLUP weights to the processed grey matter maps to create the predicted scores of MDD risks. We tested the prediction accuracy of the overall and ROI-specific BLUP predictors (that showed at least nominal significance in the UK Biobank) using logistic regression, adjusting for sex, age, BMI, smoking status and intracranial volume (ICV).

## Results

### The BLUP predictor is strongly associated with MDD risk

As part of the BLUP training, we estimated the morphometricity of the MDD status in the training cohort of UK Biobank, which corresponds to the variance explained (R^2^) by all grey-matter voxels [27]. The morphometricity was 0.061 (standard error=0.024; p-value=4.9×10^−4^), which indicated that the 415,836 voxel-wise intensity measurements accounted for 6.1% of the variance in MDD. This is higher than the morphometricity previously reported for depression score and number of depression symptoms in the UK Biobank cohort [11].

Overall, the BLUP predictor showed an AUC of 0.57 in the test cohort (483 MDD cases and 1,939 controls) (Table 1). Using the BLUP predictor, we found that MDD cases had significantly higher predicted MDD scores compared to the controls (two-sided t-test p-value=3.1×10^−5^) (Figure 1A). This difference was confirmed in both females and males (two-sided t-test p-value=0.0028 and p-value=0.0027, respectively) (Figure 1A). The accuracy of the BLUP predictor was confirmed by logistic regression, where after adjusting for covariates, every 1-SD increase in the BLUP predicted risk was significantly associated with a 28% increase in MDD risks (OR=1.28 [1.15-1.43]; p-value=1.1×10^−5^) in the mixed-sex cohort (Figure 1A). Similar associations were observed in females (OR=1.23 [1.07-1.41]; p-value=0.0035) and males (OR=1.36 [1.13-1.64]; p-value=0.001) (Figure 1A). We observed comparable OR estimates in sensitivity analyses, when further adjusting for MRI imaging covariates or antidepressant use, and when excluding individuals with a recorded MDD episode more than 5 years prior to or after MRI imaging (Supplemental Figure 2). This indicates that the BLUP predictor is sensitive to active or recent MDD events, and is especially sensitive to recent episodes occurring up to 5 years before imaging (OR=1.43 (1.18-1.73); p-value=0.00026) (Supplemental Figure 2).

**Figure 1.**
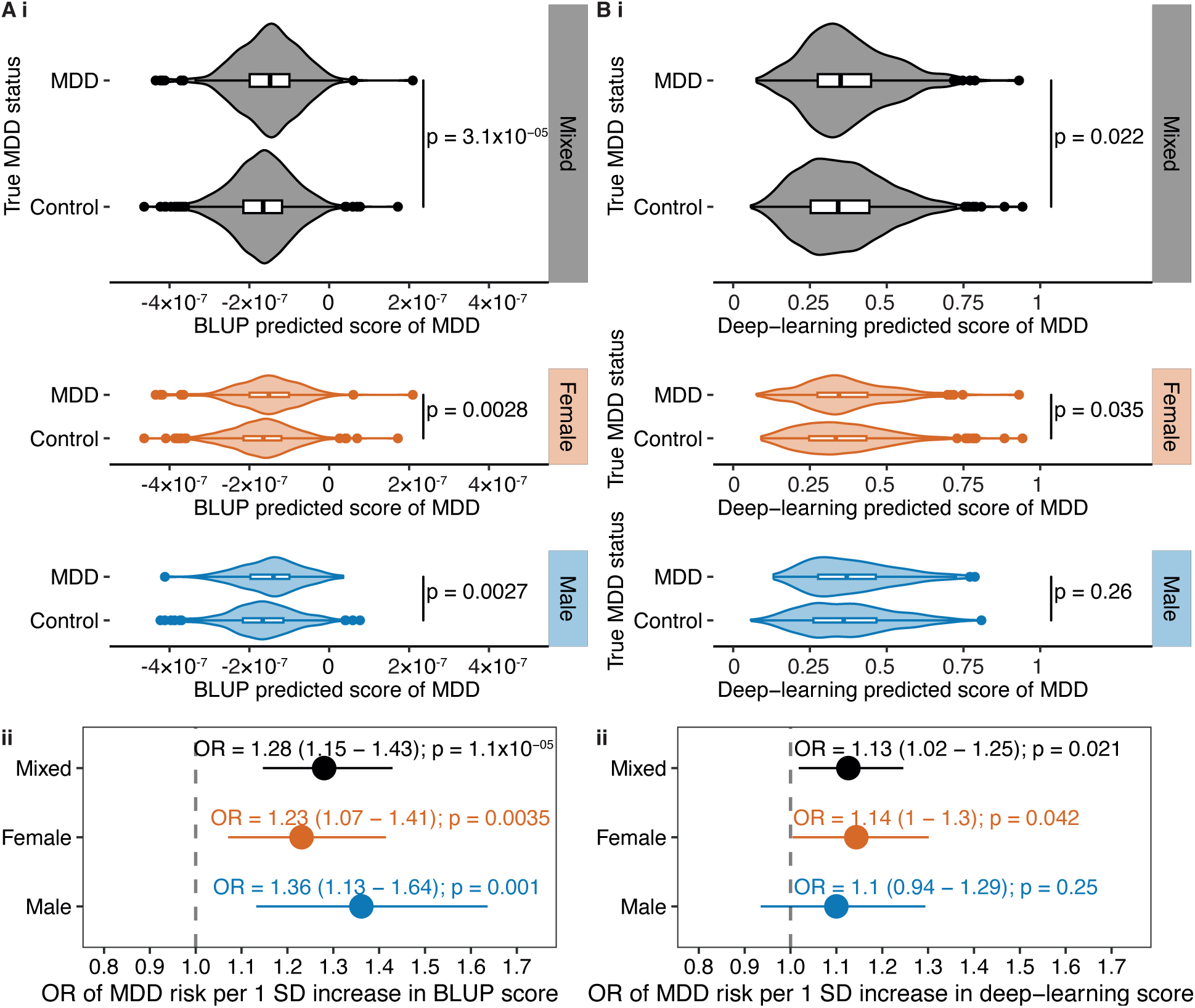
Prediction accuracy of the BLUP and deep-learning predictors in the UK Biobank test cohort. The association of the (A) BLUP and (B) deep-learning predictors with true MDD status was evaluated in the test cohort by (i) comparing the distribution of the predictors between true MDD cases and controls using t-tests, and (ii) logistic regression, adjusting for age at baseline, age at imaging, sex (except in sex-stratified analyses), genetic ancestry, smoking status at baseline, and BMI at baseline. OR represents the change in MDD risk per 1-SD increase in the BLUP or deep-learning score. The grey line represents no association (OR=1).

**Table 1.**
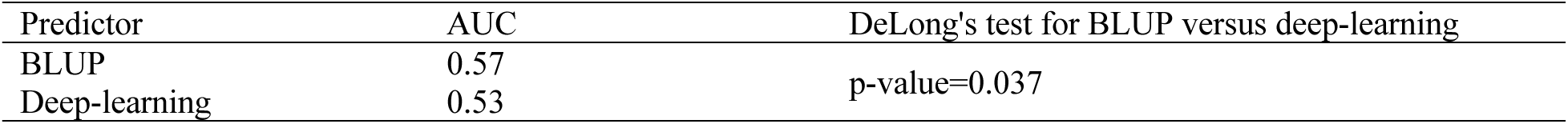
AUC of BLUP and deep-learning predictors in predicting MDD risks in the UK Biobank test cohort.

### BLUP predictor outperforms the deep-learning predictor

Our ResNet3D deep-leaning brain predictor did not show a significant association with MDD risks after multiple testing correction (Figure 1B). Overall, the deep-learning predictor exhibited an AUC of 0.53, indicating a close-to-chance capacity to distinguish MDD cases from controls (Table 1). The BLUP predictor demonstrated better performance than the deep-learning predictor (DeLong’s test p-value=0.037). To test the robustness of the deep-learning training, we trained the model using four other splits of the training and test cohorts, and found that only one split produced a deep-learning predictor with a significant association with MDD status (OR=1.16 [1.05-1.29] in the mixed-sex cohort; p-value=0.0037) (Supplemental Figure 3).

### The BLUP predictor identifies MDD-associated brain regions

To identify ROIs contributing to BLUP prediction, we partitioned the predictor based on brain anatomical regions. At p-value < 0.00033 (multiple testing correction for 150 ROIs), no ROI passed the statistical significance threshold, although 17 ROIs were nominally significantly associated with MDD risks (Figure 2). These included the left and right Crus I (cerebellum), and nine ROIs in the cortex, namely the right parahippocampal gyrus (posterior division), left parahippocampal gyrus (anterior division), left and right Heschls gyri (including H1 and H2), left and right frontal operculum cortices, right frontal medial cortex, as well as left and right central opercular cortices. We also identified six ROIs that corresponded to subcortical structures (right thalamus, left and right hippocampus, left and right amygdala as well as left cerebral white matter [which corresponded to the boundary between the left cerebral white matter and grey matter]) (Supplemental Figure 4).

**Figure 2.**
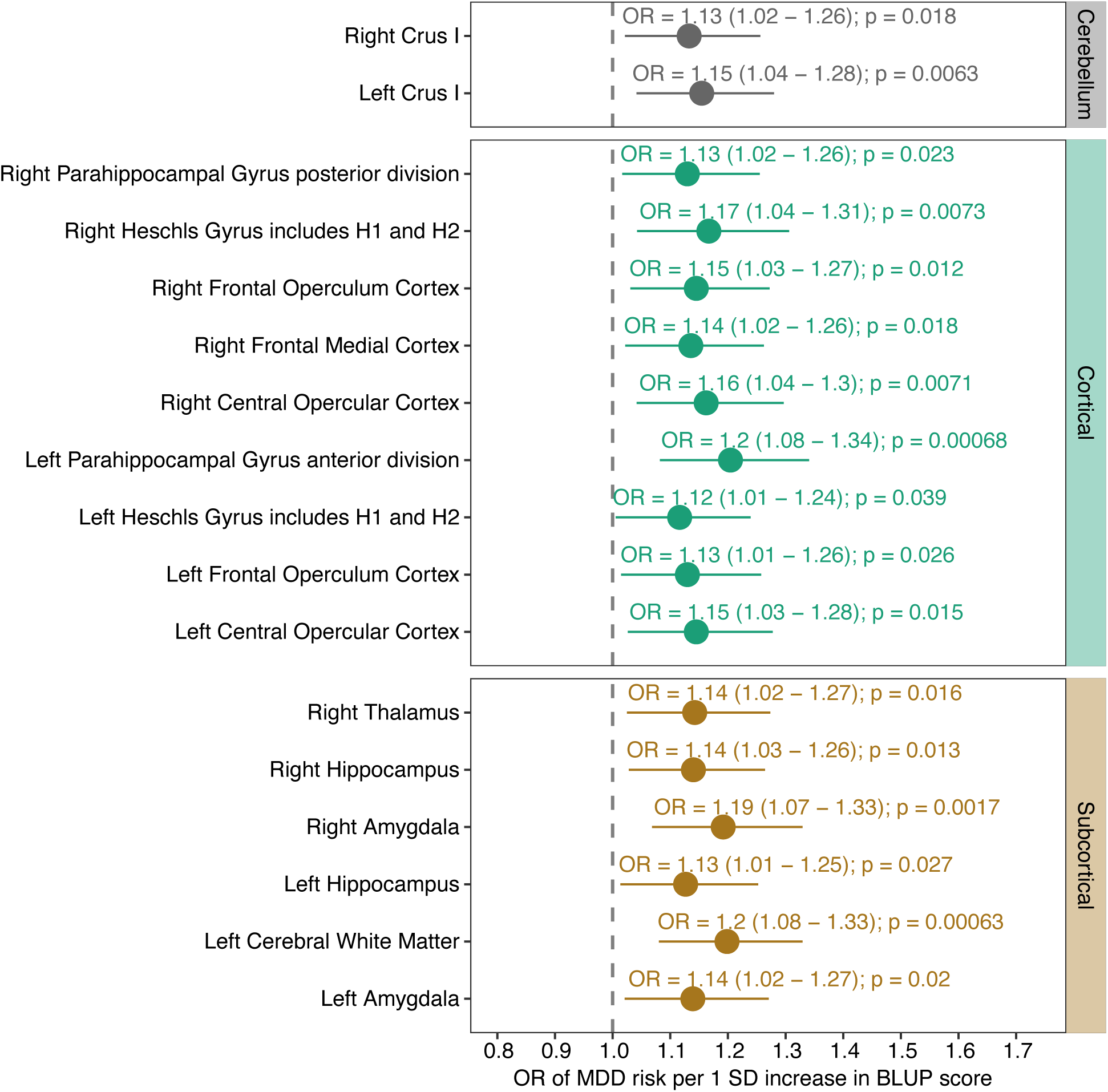
Association between MDD risks and ROI-specific BLUP predictors in the UK Biobank test cohort. Logistic regression was performed to investigate the association between each ROI-specific BLUP predictor and MDD risks. The logistic regression model included as covariates age at baseline, age at imaging, sex, genetic ancestry, smoking status at baseline, and BMI at baseline. OR represents the change in MDD risks per 1-SD increase in the BLUP score. The grey line represents no association (OR=1).

### Association with the genetically predicted risk of MDD

We found that the BLUP predictor showed comparable covariate-adjusted correlation with PGS_MD_ in the mixed and sex-stratified cohorts (indicated by similar β values), though the correlations only reached statistical significance in the mixed-sex analysis (β=0.072 [0.033-0.11]; p-value=0.0003) and the female cohorts (β=0.068 [0.019-0.12]; p-value=0.0071) (Figure 3A). The lack of statistical significance in the male cohort may be due to the reduced sample size. Nevertheless, these results suggested that the genetic and the BLUP predictor captured a common risk component of MDD. In contrast, the deep-learning predicted MDD risk was significantly correlated with PGS_MD_ in the female cohort only (β=0.068 [0.017-0.12]; p-value=0.0093), but not in the mixed-sex or male cohort (Figure 3B).

**Figure 3.**
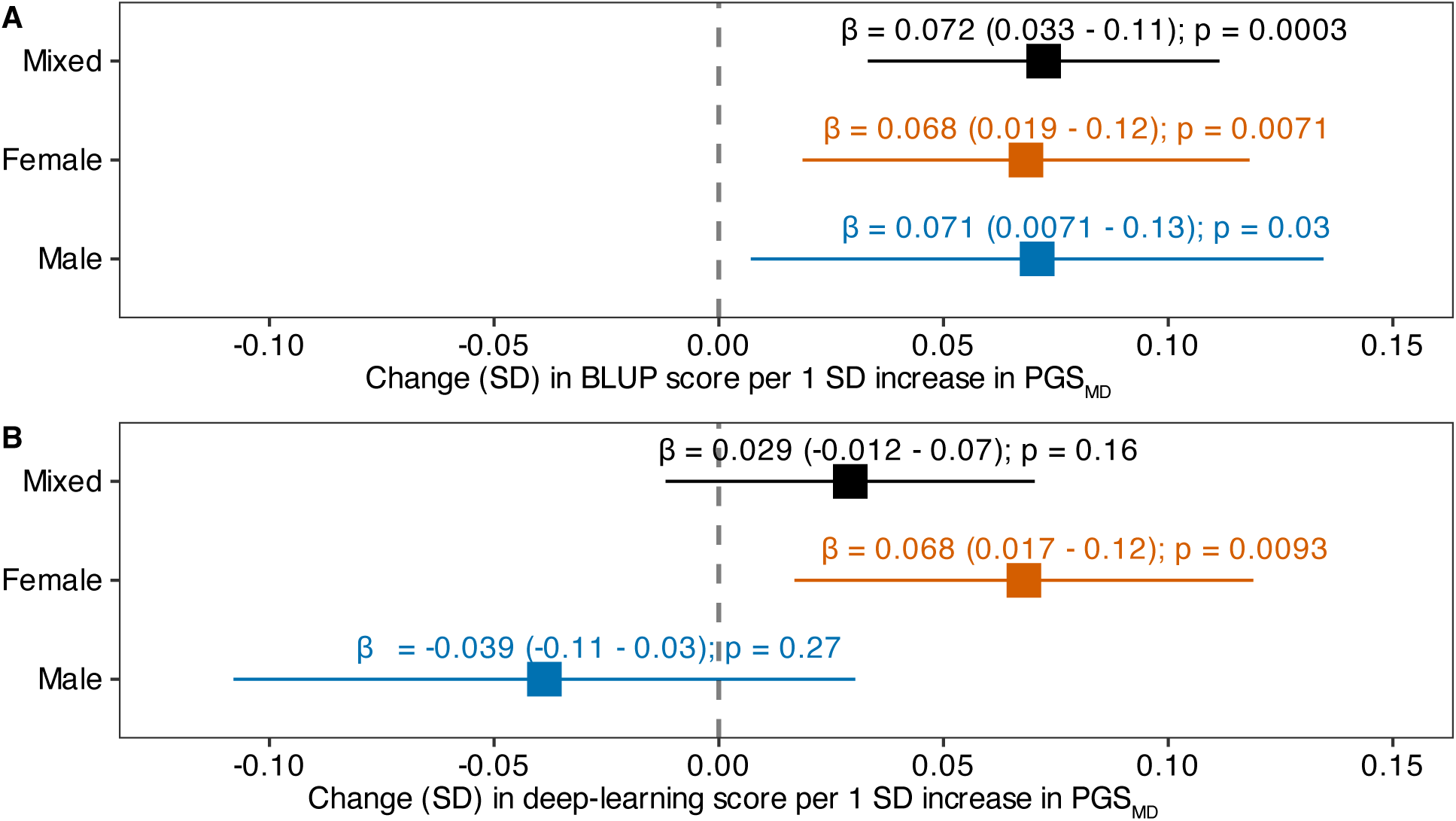
Association of PGS_MD_ with the BLUP and deep-learning scores in the UK Biobank test cohort. Linear regression was performed amongst individuals of European ancestry in the test cohort to investigate the association of PGS_MD_ with the (A) BLUP and (B) deep-learning predictors of MDD, adjusting for sex (except in sex-stratified analysis), age at baseline, age at imaging, smoking status at baseline, BMI at baseline and 10 genetic principal components. β represents the covariate-adjusted correlation between the BLUP or deep-learning predictors and PGS_MD_.

Next, we tested if combining PGS_MD_ and BLUP scores resulted in additional prediction accuracy over that of a single predictor. Amongst European-ancestry individuals in the test cohort, the PGS_MD_ alone showed an AUC of 0.65. We found that the addition of the BLUP predictor led to a modest, yet significant improvement in predicting MDD risks (AUC_PGS+BLUP_=0.66; DeLong’s test for comparison with AUC_PGS_ p-value=0.024) (Table 2). The interaction between PGS_MD_ and the BLUP predictor (PGS_MD_ * BLUP) did not improve the AUC above that of the additive model (AUC_PGS*BLUP_=0.66; DeLong’s test for comparison with AUC_PGS+BLUP_ p-value=0.61).

**Table 2.** AUC of genetic and brain-based predictors in predicting MDD risks amongst European-ancestry individuals of the UK Biobank test cohort.

| Predictor | AUC | DeLong's test for comparison with PGS<sub>MD</sub> only |
| --- | --- | --- |
| PGS<sub>MD</sub> | 0.65 | NA |
| PGS<sub>MD</sub> + BLUP | 0.66 | 0.024 |
| PGS<sub>MD</sub> * BLUP | 0.66 | 0.026 |
| PGS<sub>MD</sub> + deep-learning | 0.65 | 0.42 |

### Replication in the DEP-ARREST CLIN cohort

In the independent DEP-ARREST CLIN replication cohort (64 MDE-MDD cases and 32 controls), although the overall BLUP predictor and ROI-specific BLUP predictors were not significantly associated with MDE-MDD status in the DEP-ARREST CLIN cohort, the direction of effects for most ROI-specific BLUP predictors were concordant (i.e. positively associated with MDE-MDD) with the results from the UK Biobank cohort, including the left and right hippocampus (Supplemental Figure 5). The reduced statistical significance is most likely due to the much smaller sample size and, therefore, much limited statistical power.

## Discussion

The prediction of depression risks remains a challenging task. In this study, leveraging voxel-wise brain MRI imaging data of the grey matter, we developed a BLUP predictor that was significantly associated with MDD status (OR=1.28; AUC_BLUP_=0.57), and the prediction accuracy held up in both male and female participants. We identified 17 brain regions that were nominally significantly associated with MDD by evaluating BLUP predictors specific to each brain ROI. We found that the BLUP brain predictor contained a genetic component, although it also captured additional signals that are not currently captured by the genetic predictor, which may include brain-mediated environmental effects. In contrast, the deep-learning predictor, developed using a ResNet3D architecture, was only nominally associated with MDD risk and accounted for less variance than the BLUP predictor (OR=1.13; AUC_BLUP_=0.53). We sought to replicate the BLUP results in an independent clinical cohort (DEP-ARREST CLIN). The small sample size prevented any association from reaching significance, although the sign of most ROI-specific associations was concordant with our findings in the UK Biobank.

Previous large-consortium efforts on understanding MDD-related brain biology, such as the ENIGMA consortium [7], have identified a number of brain regions linked to MDD risks. The most consistent findings include a reduction in the hippocampus volume [2]. Furthermore, cell-type enrichment analysis of MDD-related genetic variants identified an enrichment in excitatory neurons in the midbrain, amygdala, hippocampus, thalamus and cortex [32]. Concordant with these findings, we found that the BLUP predictor specifically derived from the voxel intensities of the hippocampus was nominally significantly associated with MDD risks, further supporting its involvement in MDD aetiology. The hippocampus is a highly plastic region containing neuronal precursor cells in the dentate gyrus that give rise to new granule neurons throughout adulthood [36]. Our ROI-specific BLUP predictors also implicated brain regions whose implication in MDD remains debated, namely the amygdala and thalamus [2], as well as brain regions whose implications in MDD remain less elucidated, such as Crus I, parahippocampal gyrus, and Heschls gyrus. Overall, our findings confirm previously identified brain regions implicated in MDD and highlight previously understudied brain regions that potentially confer MDD risks and warrant further investigation.

Both genetic and environmental factors play an aetiological role in depression. Previous twin-based studies have estimated a heritability of 35%-40% for MD [37], indicating a moderate contribution of genetic factors. Although its clinical utility remains hindered by the limited prediction performance [38], PGSs, which aggregate the effects of disease-related genetic variants in the genome, have been widely used in research as a measure of lifetime risk of depression [39]. In this study, we showed that the genetically predicted MD risks were correlated with the BLUP brain predictor. However, we found that the BLUP predictor also captured some MDD risk that the latest PGS_MD_ (based on hundreds of thousands of cases) [32] does not currently tag. This could be due to the limited power of the PGS_MD_ or could reflect the brain-mediated environmental factors that contribute to depression. Several environmental risk factors have been implicated in increasing MDD risks, such as bullying, substance use, social disadvantage and personal stressful life events (e.g. divorce, financial problems and illness) [40, 41]. These risk factors have been linked to alterations in brain features that are previously implicated in MDD, such as a smaller hippocampal volume, and lower cortical thickness in the insula and the medial orbitofrontal cortex [42, 43, 44, 45]. Many of these regions overlap with or are near the brain ROIs identified in our study to contribute to the BLUP prediction. Further investigation is essential to understand how the morphological changes captured by the BLUP predictor, specifically the changes above and beyond those captured by the genetic predictor, correlate with the environmental aetiology of depression.

In our study, we found that the BLUP predictor demonstrated stronger prediction accuracy for MDD risks compared to the deep-learning predictor. Despite increasing efforts in employing deep-learning approaches in analysing brain MRI images, it was previously reported that with the current sample sizes available, deep-learning methods often performed worse than or comparably as linear or non-linear machine-learning models for prediction at the participant level [46, 47, 48]. This is in contrast with tasks where the prediction is performed at the voxel-level (for instance MRI image segmentation) and for which deep-learning methods are the state of the art, even with relatively modest sample sizes [46, 49]. This is not surprising as voxel-level supervision provides many data points of information per image for training versus only one data point for participant-level supervision.

It should be noted that although significant, the brain-based prediction accuracy of the BLUP predictor remained modest (AUC of 0.57), highlighting the difficulty of classifying MDD status from grey-matter data, and the need for a large test sample to confirm the prediction statistically. Of note, our predictor has demonstrated comparable or exceeding performance than previously reported deep-learning or machine-learning predictors trained on the ENIGMA data [8]. It is important to put in perspective that the prediction accuracy of a BLUP or linear predictor is capped by the morphometricity (which we estimated to be 6.1% of variance in the sample, corresponding to a maximum AUC of 0.64) [50]. This indicates that larger training samples could improve prediction accuracy but will never lead to a perfect predictor of MDD risk. Combining the brain and genetic predictors improved prediction, which may be useful in research, and to progress our understanding on the disease aetiology; nevertheless, these predictors combined still cannot account for the full MDD variance. This indicates that other brain modalities or measurements, such as the white matter and functional connectivity, need to be explored to further improve the prediction of MDD risk [51, 52]. In particular, one may need to focus on brain modalities that are more sensitive to environmental factors. This may be the case for functional MRI measurements, which exhibit lower heritability than structural measurements, although this may also be caused by the greater noise in measurements (lower test-retest) [53].

To our best knowledge, our study is the first to successfully develop a grey-matter predictor of MDD risks by applying machine-learning approaches to voxel-based brain MRI imaging data. Previous studies, primarily employing image-derived high-level metrics, are more computationally efficient, though at a cost of granularity. Another novelty of our study is that we demonstrated the novel use of ROI-specific BLUP scores to fine-map brain regions that most contributed to the BLUP predictor.

Limitations of our study need to be acknowledged. Firstly, the MDD cases identified in the UK Biobank consisted of a heterogenous cohort of varying symptoms, treatments, disease severity and onset ages. To address this limitation, we performed sensitivity analyses on the prediction accuracy of the BLUP predictor, where we additionally adjusted for antidepressant use or restricted the analysis to individuals with a recorded MDD episode within 5 years before or after brain imaging. At the same time, the screened controls in UK Biobank may have MDD episodes that are not in the records and thereby affect the estimated prediction accuracy. Unlike the UK Biobank, the DEP-ARREST CLIN cohort represents a more clinically homogenous cohort of MDD patients, as the cases were defined by manual adjudication by psychiatrists and were all medication-free at the time of imaging. Although the small sample size of DEP-ARREST CLIN limited the statistical power to detect any significant association, our findings showed concordant direction of associations between the UK Biobank and DEP-ARREST CLIN cohorts and warrant further replication in larger cohorts with sufficient sample sizes. Secondly, the BLUP predictor was sensitive to recent MDD episodes prior to imaging (indicated by the highest OR), though we also observed a marginal association amongst individuals with MDD episodes up to 5 years after imaging. More work is required to understand whether the brain markers underlying the BLUP predictor represent the causal or consequential effects of MDD episodes and whether the same brain markers contribute to the lifetime risk and future episodes of depression. Thirdly, substantial sex differences have been observed in the prevalence [54], heritability [37], clinical presentations [55] and neuroimaging findings [56] of MDD. Future studies with sufficient sample sizes, where the brain-based predictors are trained on female and male samples separately, are essential to provide insights into the sex-specific aetiology of depression. Lastly, brain regions previously implicated in depression, such as the hippocampus and amygdala, have also been linked to other psychiatric disorders, such as anxiety and schizophrenia [36, 57]. The utility of our BLUP predictor in informing differential diagnosis remains an outstanding yet clinically important question.

In conclusion, our study demonstrated the performance of a BLUP predictor in predicting MDD risks, and highlighted regions in the brain that contributed to the predictor. Our findings underscored the difficulty in training brain-based algorithms that can predict depression status, while suggesting promising avenues as more data become available. By integrating genetic information, we showed that our BLUP brain scores likely reflect the joint effects of genetic and environmental risk factors on depression. Future studies on the development of symptom-specific predictors will further enrich the current knowledge on the function and pathophysiological links of specific brain regions in MDD. Furthermore, longitudinal studies with medication and treatment information available (such as DEP-ARREST CLIN) are essential for understanding the mechanisms by which pharmacological treatments of depression reverse or improve disease symptoms.

## Data Availability

All data produced in the present study are available upon reasonable request to the authors.

## Acknowledgements

Sonia Shah is supported by a Heart Foundation Fellowship. Funding for the DEP-ARREST-CLIN cohort, where Emmanuelle Corruble is the principal investigator, was provided by a national grant (ANR SAMENTA 2012) of the Agence Nationale de la Recherche (ANR) and sponsored by the Institut National de la Santé et de la Recherche Médicale (INSERM) (C13-25, EudraCT 2013-004326-29). The research contributing to these results has received funding from the French government under management of Agence Nationale de la Recherche as part of the “France 2030” program (reference ANR-23-IACL-0008, project PRAIRIE-PSAI), as part of the “Investissements d’avenir” program (reference ANR-19-P3IA-0001, project PRAIRIE 3IA Institute and reference ANR-10-IAIHU-06, project Agence Nationale de la Recherche-10-IA Institut Hospitalo-Universitaire-6), from the European Union’s Horizon Europe Framework Programme (grant number 101136607, project CLARA) and by Inria in the context of the Inria-University of Queensland international team (“Brainetics”). Baptiste Couvy-Duchesne is supported by Inria (starting faculty position) and an NHMRC CJ Martin Fellowship (app 1161356).

## Participant consent statement and ethics approval

Informed consent was obtained from all UK Biobank participants by the UK Biobank. Procedures are controlled by a dedicated Ethics and Guidance Council (http://www.ukbiobank.ac.uk/ethics), with the Ethics and Governance Framework available at https://www.ukbiobank.ac.uk/media/0xsbmfmw/egf.pdf. IRB approval was also obtained from the North West Multi-centre Research Ethics Committee. Our research has been conducted using the UK Biobank Resource under Application Number 12505. This research is covered by The University of Queensland Human Research Ethics Committee approval (HREC number 2020/HE002938).

Written informed consent of the DEP-ARREST CLIN participants was obtained after the nature of the procedures had been fully explained.

## Supplemental Figures

**Supplemental Figure 1.**
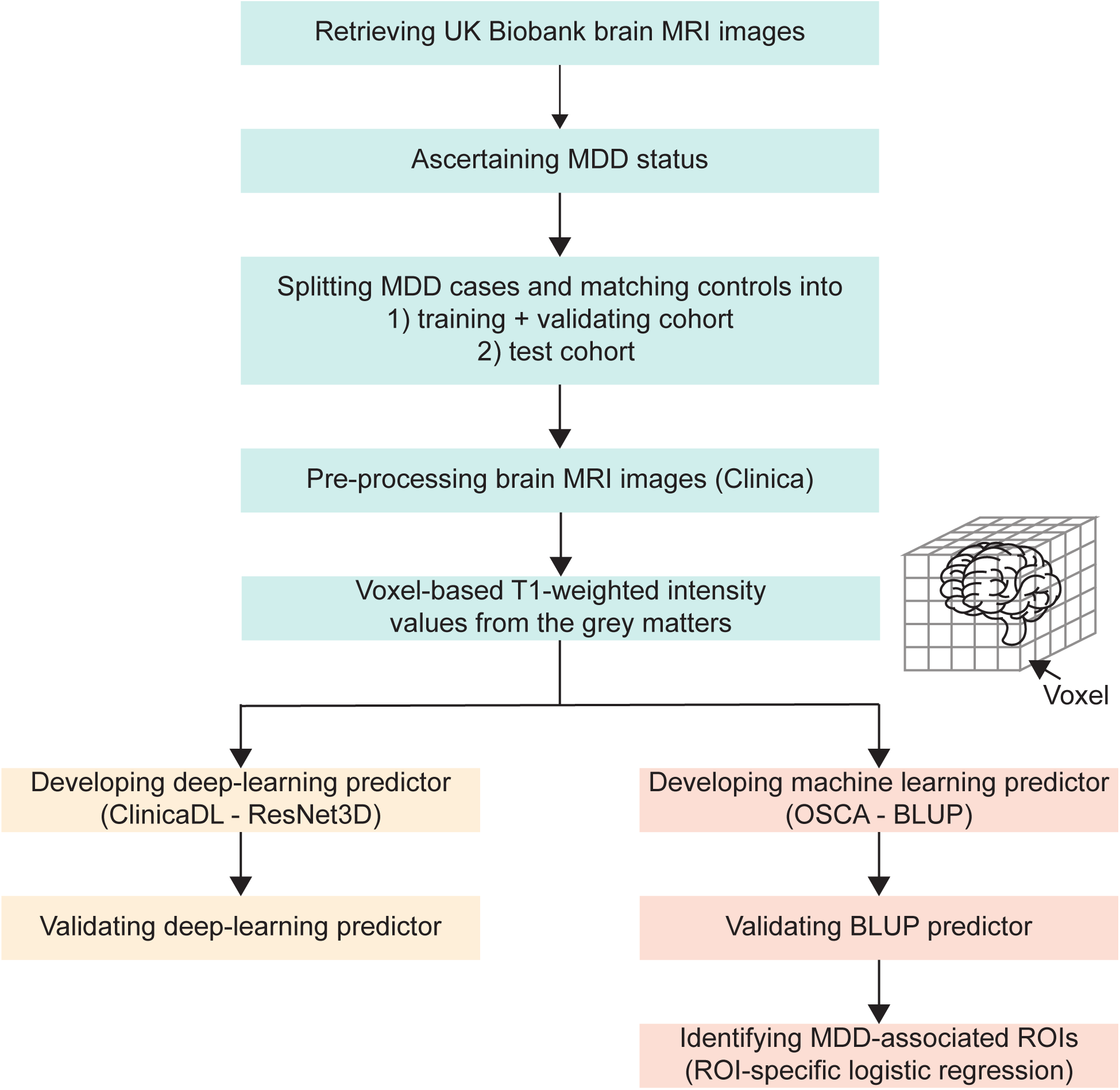
Overview of study design.

**Supplemental Figure 2.**
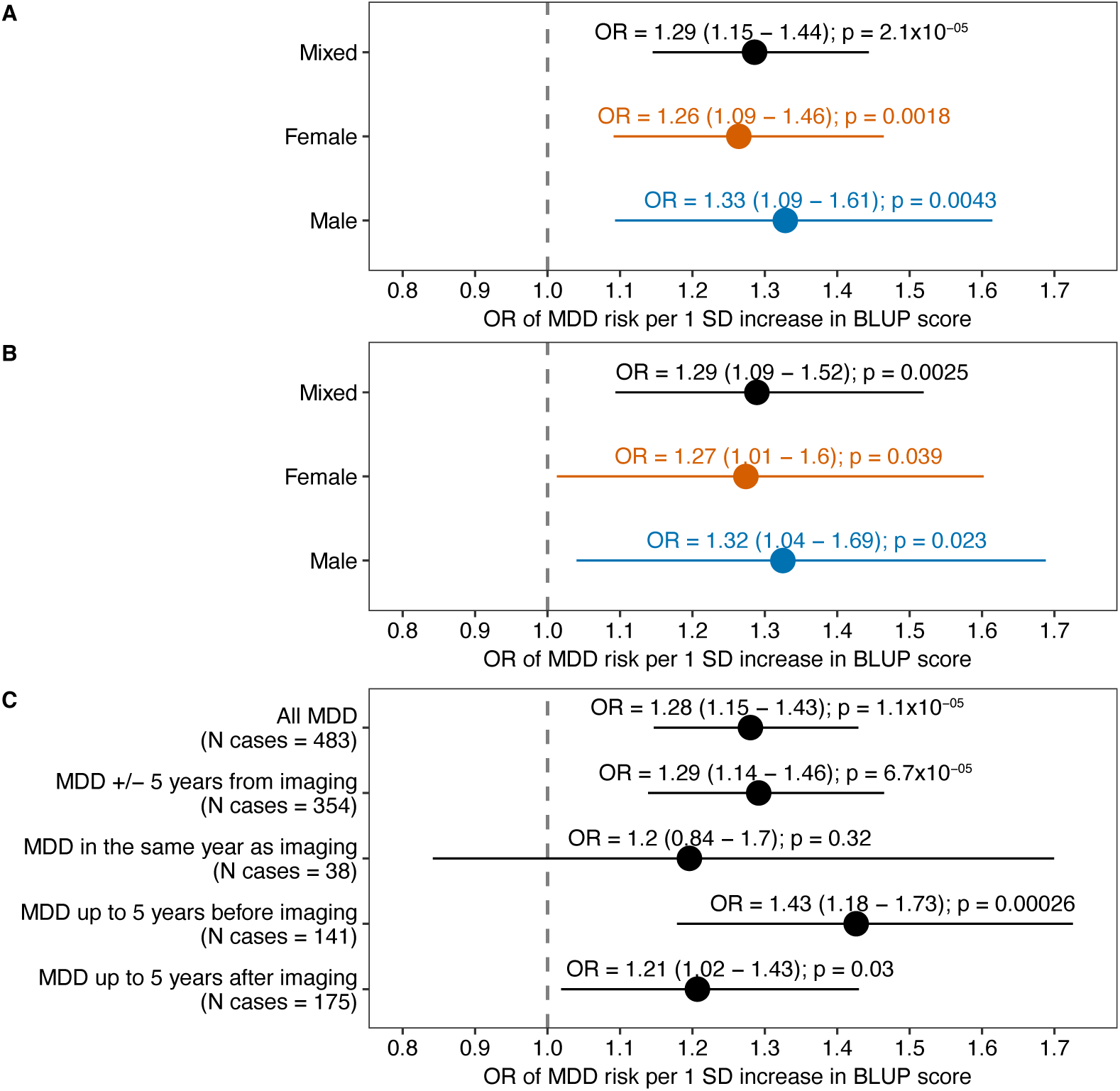
Sensitivity analyses of the association between the BLUP predictor and MDD risks. Logistic regression was performed to investigate the association between the BLUP predictor and MDD risks, while adjusting for age at baseline, age at imaging, sex (except in sex-stratified analyses), genetic ancestry, smoking status at baseline, and BMI at baseline. (A) Logistic regression was performed while additionally adjusting for MRI imaging covariates, namely mean rfMRI head motion, scanner lateral (X) brain position, scanner transverse (Y) brain position, scanner longitudinal (Z) brain position, intensity scaling for T1, volume of EstimatedTotalIntraCranial (whole brain), inverted signal-to-noise ratio in T1, and inverted contrast-to-noise ratio in T1. (B) Logistic regression was performed while additionally adjusting for antidepressant use. (C) Logistic regression was performed where MDD cases with a recorded MDD episode more than 5 years prior to or after MRI imaging were excluded from analysis. Logistic regression was adjusted for age at baseline, age at imaging, sex, genetic ancestry, smoking status at baseline, and BMI at baseline. OR represents the change in MDD risks per 1-SD increase in the BLUP score. The grey line represents no association (OR=1).

**Supplemental Figure 3.**
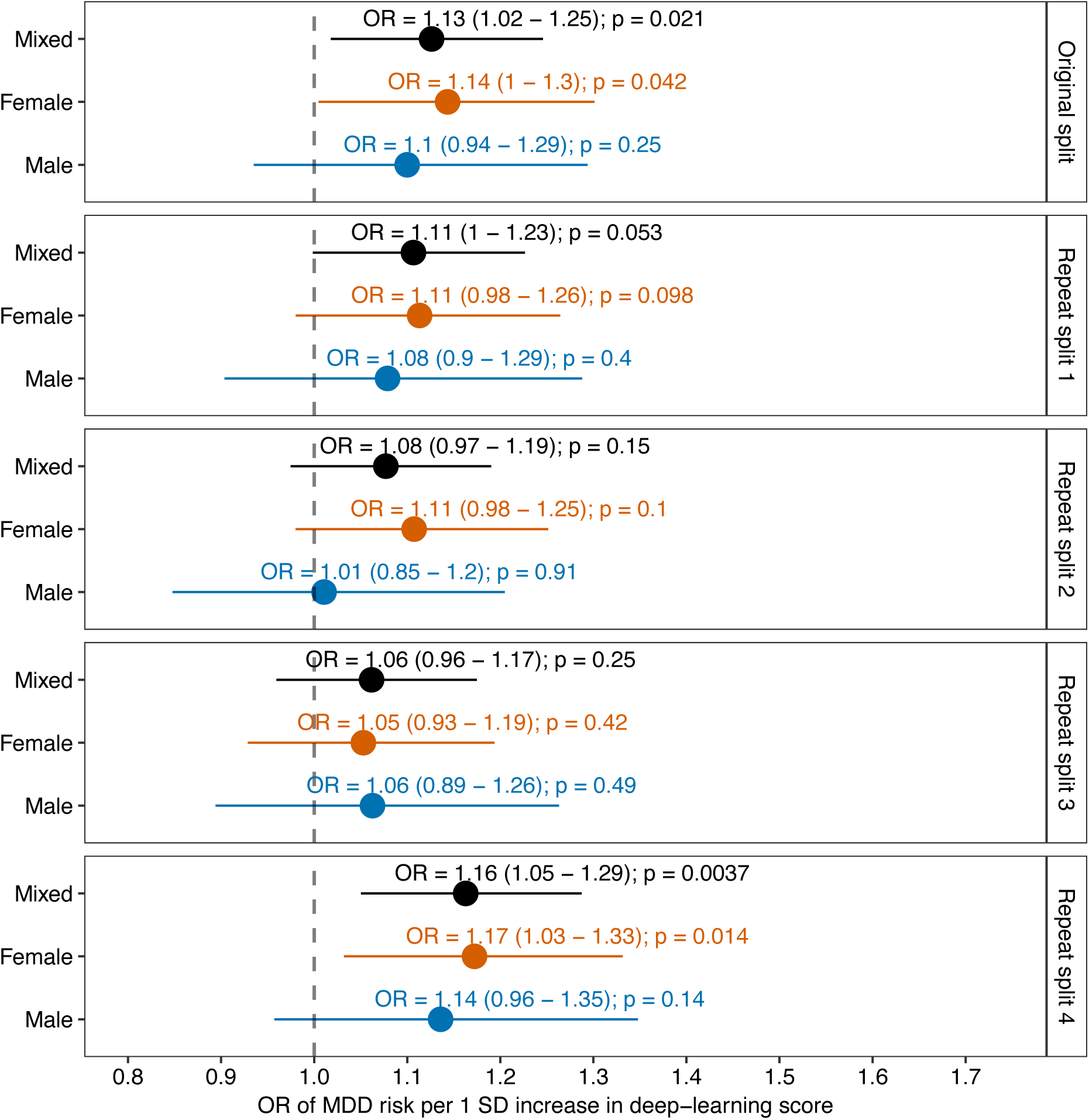
Association between MDD risks and deep-learning predictors developed in the repeated splits of the training cohort. Logistic regression was performed to investigate the association between the deep-learning predictor and MDD risks. The logistic regression model included age at baseline, age at imaging, sex (except in sex-stratified analyses), genetic ancestry, smoking status at baseline, and BMI at baseline as covariates. OR represents the change in MDD risks per 1-SD increase in the deep-learning score. The grey line represents no association (OR=1).

**Supplemental Figure 4.**
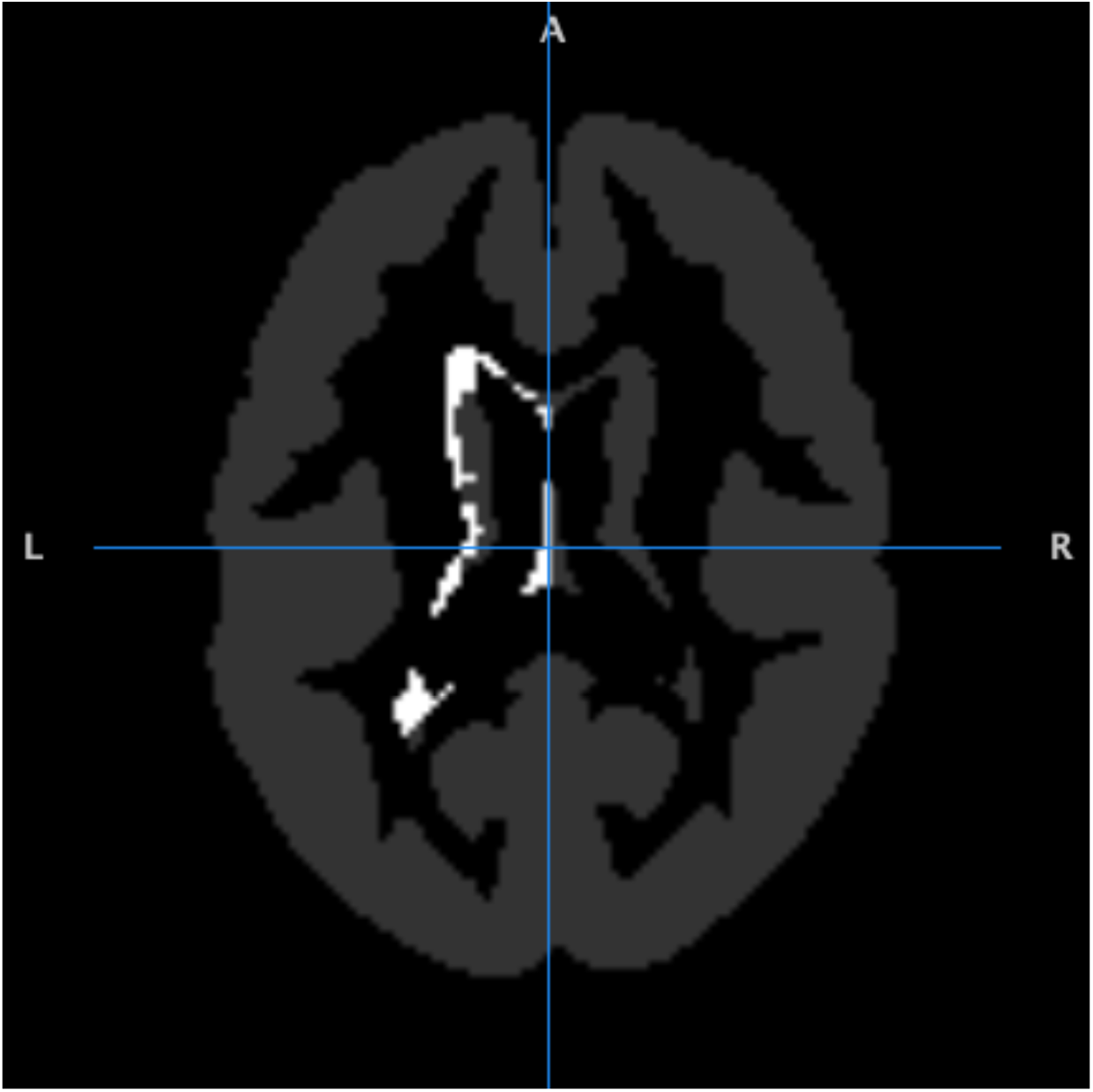
Voxels mapping to the left cerebral white matter. Black indicates voxels that were not annotated with a grey matter region of interest. Grey indicates voxels that were annotated with a region of interest. White indicates voxels that map to the left cerebral white matter.

**Supplemental Figure 5.**
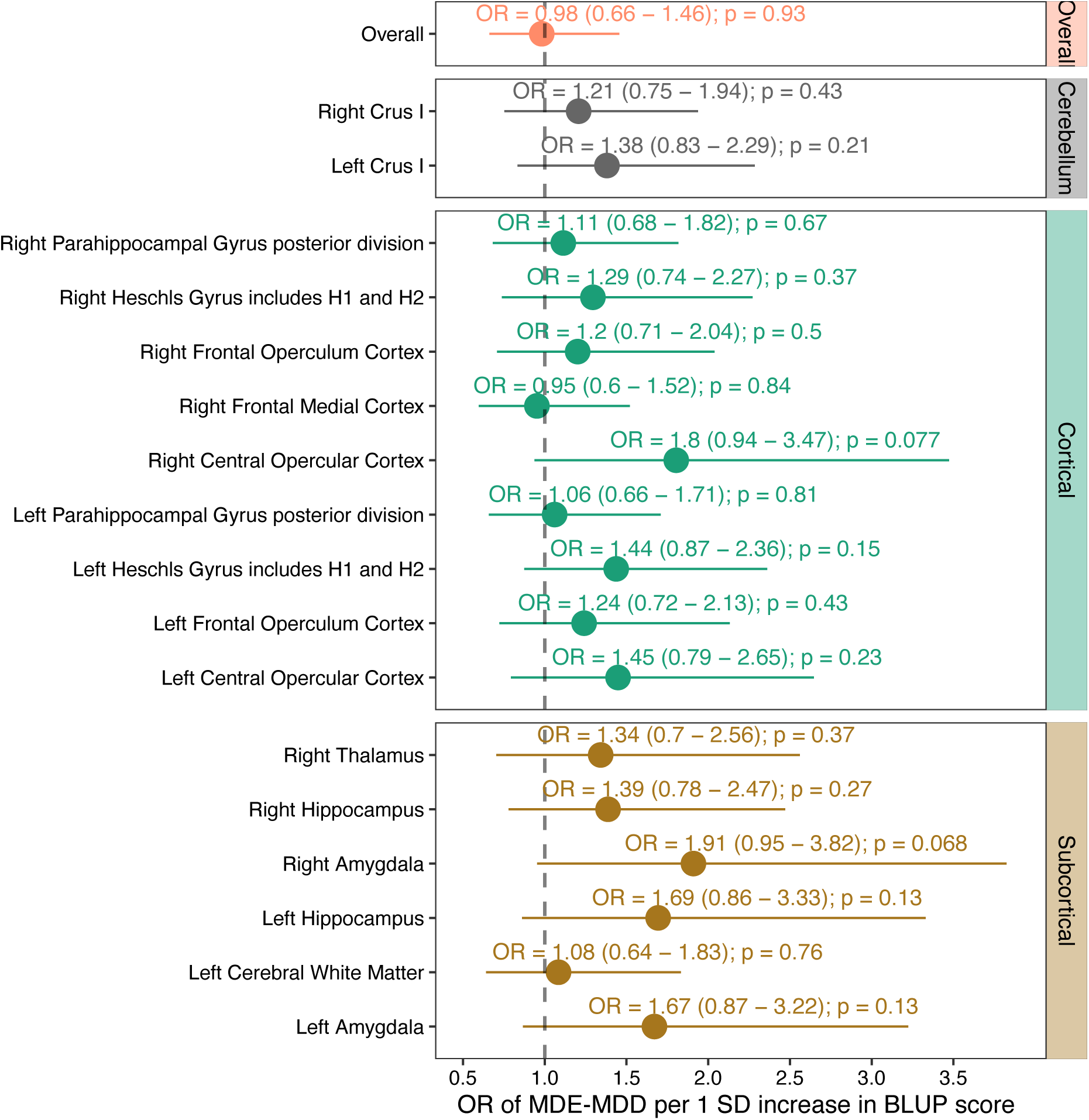
Association between MDE-MDD status and the overall and ROI-specific BLUP predictors in the DEP-ARREST CLIN replication cohort. Logistic regression was performed to investigate the association between each BLUP predictor and MDE-MDD status. The logistic regression model included age, sex, BMI, smoking status and ICV as covariates. OR represents the change in MDE-MDD per 1-SD increase in the BLUP score. The grey line represents no association (OR=1).

## Supplemental Tables

**Supplemental Table 1.**
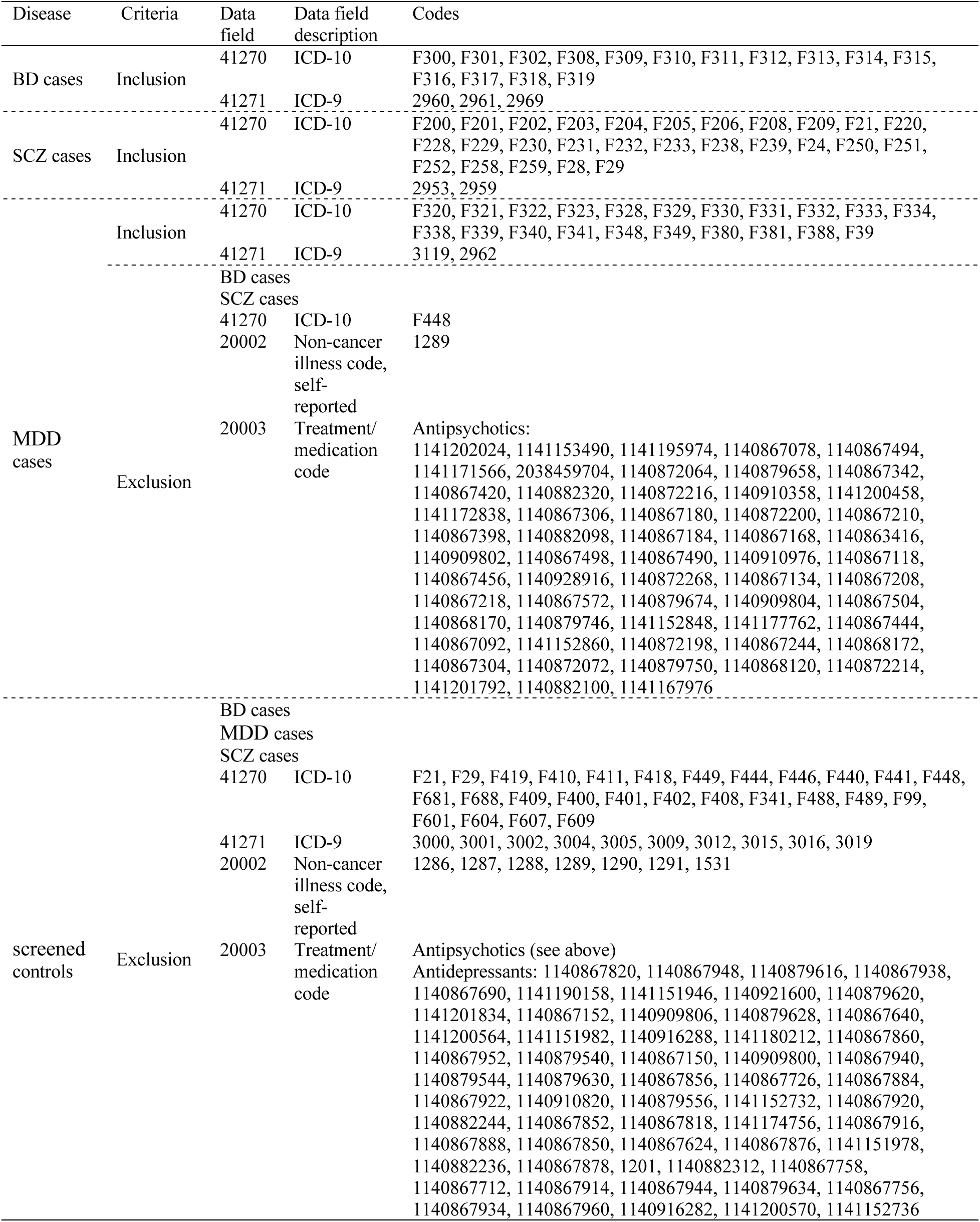
UK Biobank MDD ascertainment criteria (BD, bipolar disorder; SCZ, schizophrenia)

**Supplemental Table 2.**
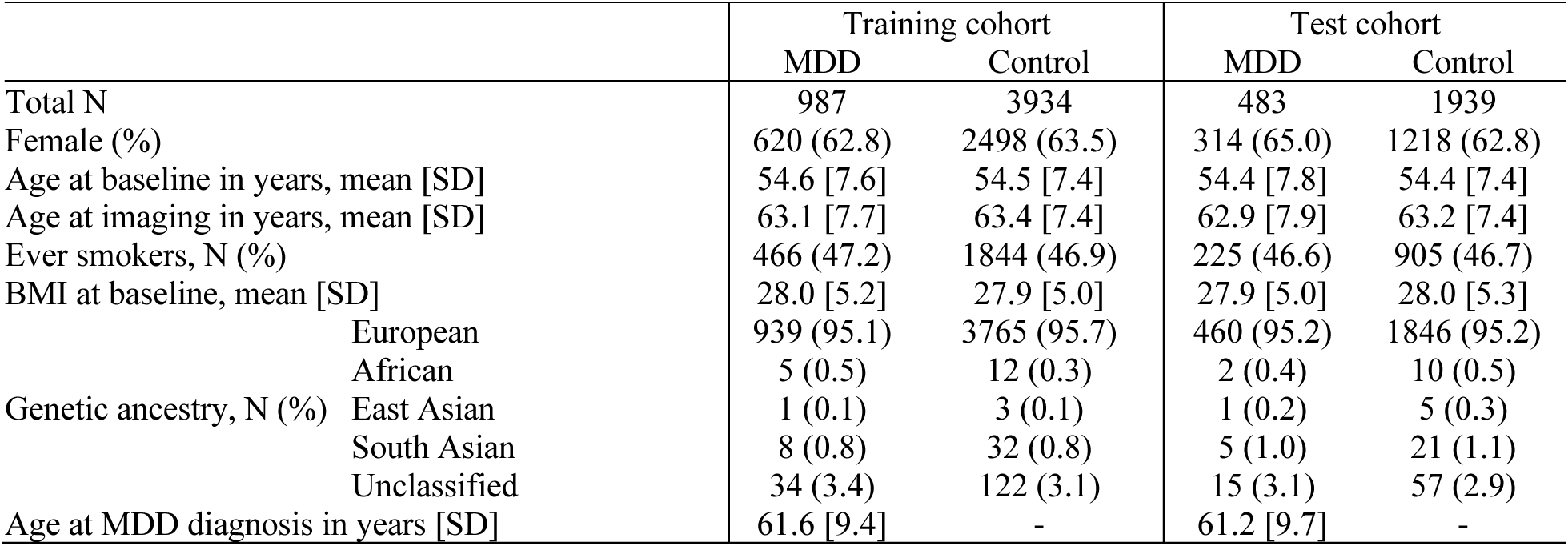
Demographic characteristics of the training and test cohorts.

